# Microstructural changes in the reward system are associated with post-stroke depression

**DOI:** 10.1101/2020.01.14.20017384

**Authors:** Lena KL Oestreich, Paul Wright, Michael J O’Sullivan

## Abstract

**Background:** Studies of lesion location have been unsuccessful in identifying simple mappings between single brain regions and post-stroke depression (PSD). This might partly reflect the involvement of multiple interconnected regions in the regulation of mood. In this study, we set out to investigate whole-brain network structure and white matter connectivity in the genesis of PSD. Based on studies implicating regions of the reward system in major depressive disorder without stroke, we investigated the overlap of whole-brain correlates of PSD with this system and performed a focused analysis of grey matter and white matter projections within the reward system and their associations with the development of PSD.

**Methods:** The study enrolled 46 patients with first ischemic stroke, 12 were found to have PSD (D+ group) and 34 were free of PSD (D-) based on scores on the Geriatric Depression Scale. A group of 16 healthy controls were also recruited. Participants underwent research MRI with 3T structural and diffusion sequences. Graph theoretical measures derived from measures of microstructure were used to examine global topology and whole-brain connectome analyses were employed to assess differences in the interregional connectivity matrix between the three groups. Structural correlates specific to the reward system were examined by measuring grey matter volumes from regions in this circuit and by reconstructing its main white matter pathways, namely the medial forebrain bundle and connections within the cingulum bundle with deterministic tractography. For network connections and tracts, we derived measures of microstructural organization (FA), and also extracellular free-water content (FW) as a possible proxy of neuroinflammation.

**Results:** The topology of structural networks differed across the three groups. Network modularity, weighted by extracellular FW content, increased with depression severity and connectome analysis identified networks of decreased FA-weighted and increased FW-weighted connectivity in patients with PSD relative to healthy controls. Intrinsic frontal and fronto-subcortical connections were a notable feature of these networks, which also subsumed the majority of regions defined as constituting the reward system. Within the reward system, grey matter volume of cortical and subcortical regions, as well as FA and FW of major connection pathways, were collectively predictive of PSD severity, explaining 76.8% of the variance in depression severity.

**Conclusions:** Taken together, these findings indicate that PSD is associated with microstructural characteristics of the reward system, similar to those observed in major depressive disorder without stroke. Alterations in the reward system appear to drive differences in whole-brain network structure found in patients with PSD. Even in the absence of a simple relationship with lesion size and location, neuroimaging measures can explain much of the variance in depression scores. Structural characterization of the reward system is a promising biomarker of vulnerability to depression after stroke.

## Introduction

Post-stroke depression (PSD) is a common complication after stroke, with approximately 31% of stroke survivors meeting the criteria for major depression 3-6 months after stroke.^1^ Patients with PSD have increased disability, mortality and poorer rehabilitation outcomes, compared with individuals free of depression.^2^ Despite these well-known, detrimental effects on functional recovery, recognition and treatment of PSD remains suboptimal.^2^ A possible factor is a poor understanding of the underlying brain-based biological mechanisms. Studies that have investigated associations between lesion location and PSD have generated inconsistent and often contradictory findings, leaving the field unable to reach a consensus for the mechanistic basis of PSD.^3^ Early qualitative approaches based on visual inspection to determine lesion location reported higher incidences of PSD in patients with left hemisphere lesions,^4,5^ but were soon complemented by studies showing the opposite pattern, with an association between PSD and right hemisphere lesions.^6,7^ More recent studies using voxel-based lesion symptom mapping, which normalizes and co-registered brain imaging data into a standard template and therefore represents a more quantitative approach to study lesion locations, have also reported conflicting findings.^8-10^ This inconsistency among single studies is reflected in systematic reviews^3,11,12^ and meta-analyses,^13,14^ which have been unable to reveal any associations between lesion locations and PSD.

The inability to pinpoint lesion locations specific to PSD has led some to question whether PSD might arise from more diffusely distributed pathogenic mechanisms, such as widespread activation of inflammatory mechanisms. Systemic inflammation has recurrently been implicated in major depressive disorder (MDD).^15^ However, accumulating evidence suggests that even in the presence of systemic mechanisms, the causative alterations reside in relatively circumscribed brain regions.^16^ Structural connectome studies reported disrupted white matter connectivity^17,18^ primarily localized to subcortical-frontal regions in MDD. Furthermore, a connectome study in PSD reported impaired network integration and segregation in fronto-limbic regions to be associated with depression severity.^19^ These brain circuits are commonly referred to as the reward system, which is essential for emotional and motivational information processing and plays a pivotal role in memory.^16^ It consists of subcortical and fronto-cortical regions,^16^ which are interconnected by white matter projections of the cingulum and medial forebrain bundle. Multiple neuroimaging studies have provided evidence for grey matter volume reductions in the reward system^20,21^ and microstructural changes in the medial forebrain bundle^22,23^ and the cingulum bundle^23,24^ to be implicated in MDD.

Based on the lack of consistent lesion locations associated with PSD, together with mounting evidence for structural changes in the reward system associated with MDD, we set out to investigate network level and reward system structural features as substrates of PSD. Structural connectome analyses and global topology were used to assess connectivity differences between stroke patients with and without PSD, and healthy controls. We then investigated structural changes specifically localized to the reward system by reconstructing its main white matter pathways and parcellating its main grey matter structures. We hypothesized that patients with PSD would exhibit connectivity disruptions relative to healthy controls and stroke patients without PSD particularly in structures constituting the reward circuit. We furthermore hypothesized that depressive symptom severity would beassociated with microstructural alterations in patients with recent stroke.

## Methods

### Participants

Participants ranged in age from 51 to 86 years (*M* = 70.04, *SD* = 9.07), 37.1% (*n* = 23) were female, and 97% (*n* = 60) were right-handed (see Table 1). Patients with first ischemic stroke were enrolled into a longitudinal study (STRATEGIC) within 7 days of stroke. Inclusion criteria were age over 50 years and clinical stroke confirmed by CT or MRI. Exclusion criteria were dementia, previous stroke, inability to converse fluently in English, major neurological disease, active malignancy, previous moderate to severe head injury and any other factor that would prevent performance of cognitive tasks (e.g. visual impairment). The study procedures were approved by the London and Bromley Research Ethics Committee and the University of Queensland Research Ethics Committee. All participants gave written informed consent. Forty-six out of 179 participants were enrolled in an in-depth substudy. These individuals completed the Geriatric Depression Scale (GDS), a 30-item self-report measure to identify depression in older people^25^ 27-82 days after stroke (*M* = 42.95, *SD* = 13.95) and underwent MRI 30-95 days after stroke (*M* = 65.76, *SD* = 17.16). Sixteen healthy controls were recruited from the community (see Table 1). Thirty-four (73.9%) stroke patients had a score below 10 on the GDS and were assigned to the group without PSD (D-). The remaining 12 (26.1%) participants scored 10 or above on the GDS and were assigned to the PSD group (D+).

**Table 1.** Demographics, risk factors and lesion characteristics by group

|  |  |  |  |  |
| --- | --- | --- | --- | --- |
|  | HC (*n* = 16) | D+ (*n* = 12) | D- (*n* = 34) |  |
| Variable | Mean (*SD*)/ category | Mean (*SD*)/ category | Mean (*SD*)/ category | Group comparisons |
| *Demographics* |  |  |  |  |
| Age (years) | 71.53 (10.62) | 69.72 (6.96) | 69.46 (9.13) | $F(2,59) = 0.29, p = 0.75$ |
| Sex (female/male) | 11/5 | 3/9 | 9/25 | $\chi^2(2) = 9.27, p = 0.01$ |
| Handedness (right/left) | 16/0 | 11/1 | 33/1 | $\chi^2(2) = 1.55, p = 0.47$ |
| *Risk factors* |  |  |  |  |
| ECG (normal/sinus rhythm/atrial fibrillation) | | 1/10/1 | 1/23/10 | $\chi^2(2) = 2.55, p = 0.28$ |
| Hypertension (no/yes<sup>a</sup>/yes<sup>b</sup>) | | 5/7/0 | 15/15/4 | $\chi^2(2) = 1.8, p = 0.41$ |
| Diabetes mellitus (no/yes<sup>c</sup>/yes<sup>d</sup>/yes<sup>e</sup>/yes<sup>f</sup>) | | 10/0/1/1 | 28/3/3/0 | $\chi^2(3) = 3.9, p = 0.27$ |
| Smoking (never/previously/current) | | 5/6/1 | 19/10/5 | $\chi^2(2) = 1.7, p = 0.43$ |
| Ischemic heart disease (no/yes) | | 10/2 | 27/7 | $\chi^2(1) = 0.09, p = 0.77$ |
| Statins (no/yes<sup>g</sup>/yes<sup>h</sup>) | | 1/7/4 | 2/21/11 | $\chi^2(2) = 0.1, p = 0.95$ |
| *Lesion characteristics* |  |  |  |  |
| Hemisphere (left/right) | | 5/7 | 19/15 | $\chi^2(1) = 0.72, p = 0.4$ |
| Arterial territory (MCA<sub>ant</sub>/MCA<sub>pos</sub>/<br>MCA<sub>str</sub>/PCA/lacunar/thalamic) | | 2/3/3/2/1/1 | 7/7/5/8/5/2 | $\chi^2(5) = 1.24, p = 0.94$ |
| Volume (ml) | | 7175.64 (12081.03) | 7947.09 (11917.81) | $t(44) = 0.73, p = 0.47$ |
| Time since lesion (days) | | 68.17 (13.39) | 79.88 (54.63) | $t(44) = 0.19, p = 0.85$ |

*Note.* HC = healthy controls; D+ = Depression group; D- = no Depression group; SD = standard deviation; ECG = electrocardiogram;
<sup>a</sup>controlled, <sup>b</sup>uncontrolled; <sup>c</sup>type 1, <sup>d</sup>type 2 controlled by diet, <sup>e</sup>type 2 controlled by tablets, <sup>f</sup>type 2 controlled by insulin injections; <sup>g</sup>normal lipids,
<sup>h</sup>abnormal lipids; MCA<sub>ant</sub> = middle cerebral artery, anterior; MCA<sub>pos</sub> = middle cerebral artery, posterior; MCA<sub>str</sub> = middle cerebral artery, striatocapsular; PCA = posterior cerebral artery

Detailed information on data acquisition, pre-processing, and analysis are provided in the Supplements.

### Data acquisition

MRI scans were collected on a 3T MR750 MR scanner (GE Healthcare, Little Chalfont, Buckinghamshire, UK). T1-weighted images were acquired with the MPRAGE sequence^26^ and diffusion-weighted images with an echo planar imaging sequence with double refocused spin echo for 60 diffusion-sensitization directions at *b*=1500s/mm^2^ and six acquisitions without diffusion sensitization (*b*=0).

### Connectome reconstruction

Cortical and subcortical parcellations were reconstructed from the T1-weighted images, based on the Desikan-Killiany atlas, resulting in 84 connectome nodes.^27^ The diffusion-weighted data were pre-processed using tools implemented in MRtrix3^28^ to correct for head movements, eddy current distortions and field inhomogeneities. Individual tractograms were reconstructed and connectivity matrices were generated by mapping streamlines onto nodes of each participant’s parcellation image. Separate connectivity matrices were populated with fractional anisotropy (FA) and free-water (FW).

### Graph theoretical measures

The following global network metrics were calculated on the interregional connectivity matrices: global efficiency, which estimates the overall integration of a network, modularity, a metric of a network’s segregation into multiple subnetworks, and average global clustering coefficient, which quantifies the connectivity strength of all closed triangles a node forms with its neighbouring nodes.

### Measurements from the reward system

Grey matter regions of the reward system were selected based on a literature review of fMRI and structural MRI studies of the reward system in depression (see Table S1). Grey matter volumes were calculated for the amygdala, nucleus accumbens, thalamus, hippocampus, caudate, putamen, dorsolateral prefrontal cortex, medial prefrontal cortex, orbitofrontal cortex, anterior cingulate cortex and insula. The cingulum bundle and medial forebrain bundle (MFB), were reconstructed with deterministic tractography. The cingulum bundle was divided into anterior, middle, posterior and parahippocampal subdivisions and the MFB was reconstructed as a single tract in each hemisphere. FA and FW were averaged across each tract.

### Statistical analysis

Voxel-based symptom lesion mapping (VSLM) was performed on the co-registered lesion images with lesion location as independent variable and GDS scores as outcome variable. Global topological group differences were tested for the global graph metrics *global efficiency (FA/FW), modularity (FA/FW)*, and *centrality coefficient (FA/FW)* and network-based statistics (NBS) were used to investigate whole-brain between-group differences in FA and FW.^29^ Focused analyses on measures within the rewards system were conducted by investigating group differences on FA and FW in the MFB and cingulum subdivisions as well as the grey matter volumes. In order to investigate whether global graph theoretical metrics or structural changes in the reward system account for PSD severity, two multiple linear regression analysis were performed in the stroke sample only.

## Results

The three groups did not differ significantly on age or handedness, but there were more women among the healthy controls and more men in the stroke groups but a similar gender ratio (approximately 1:3 women to men) in both D+ and D-groups (see Table 1). The D+ and D-groups did not differ significantly on any measures of vascular risk factors or lesion characteristics (see Table1).

### Associations between depression and lesion characteristics

VLSM identified a small cluster of 1195 voxels in the left putamen and part of the MFB (see Figure S1C) associated with GDS scores but the strength of association was modest (*z* = 2.438, *p*_*uncorr*_ = 0.008).

### Whole-brain topology and connectome analysis

There was a significant main effect of *group* on *modularity FW* (*F*(3,61) = 2.239, *p* = 0.046,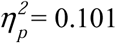). *Modularity FW* significantly increased linearly from the HC group, to the D- and the D+ group (*t*(59) = 2.394, *p* = 0.02). The D+ group had significantly increased *modularity FW* (*t*(59) = 2.382, *p*_*corr*_ = 0.045) compared to the HC group.

Compared to the HC group, the D+ group showed reduced FA-weighted connectivity (*p*_*FWE*_ = 0.029) in a subnetwork that subsumed 77% of nodes located in the reward system. Frontal-subcortical and within frontal lobe connections were a notable anatomical feature of this subnetwork. A subnetwork of increased FW-weighted connectivity in the D+ group compared to the HC group (*p*_*FWE*_ = 0.038) was also demonstrated. This subnetwork also demonstrated an emphasis on connections (edges) to and within the frontal lobes (62%) and included 80% of nodes from the reward system (see Figure 1).

**Figure 1.**
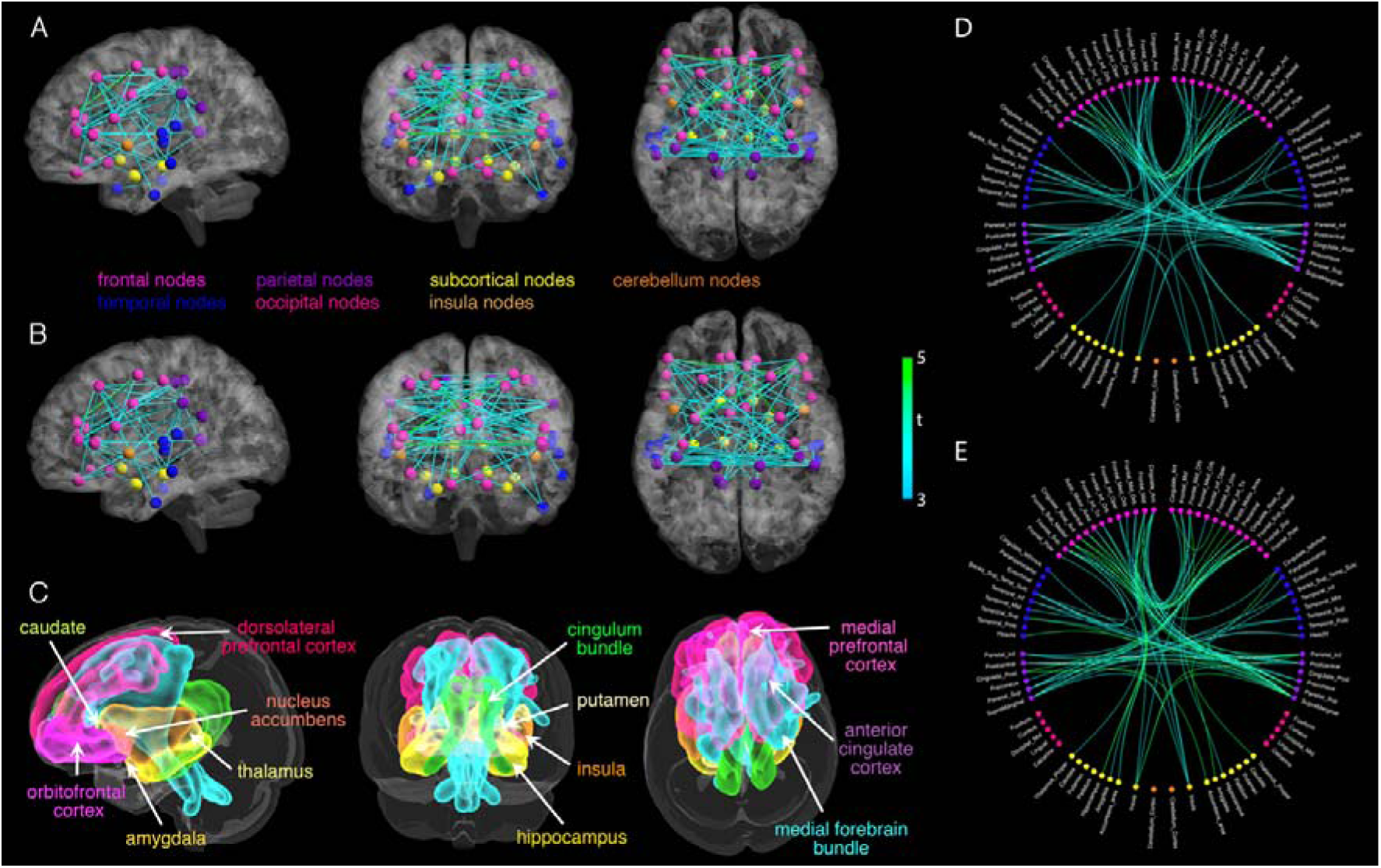
A) Network of significantly reduced fractional anisotropy (FA)-weighted connectivity in the group of stroke patients with depression compared to the healthy control group. B) Network of significantly increased free-water (FW)-weighted connectivity in the group of stroke patients with depression compared to the healthy control group. T-statistics are set to a supra-threshold of 3, which corresponds to *p* = 0.001. Subnetworks are significant at p_FWE_ < 0.05. C) Grey matter (warm colors) and white matter (cold colors) structures constituting the reward system. The medial forebrain bundle (cyan) and the cingulum bundle (green) interconnect the grey matter structures of the reward system. Connectograms of the significant D) FA and E) FW networks. Edge color correspond to nodes in different lobes and subcortical regions. Green color represents higher F-statistics.

### Structural group differences in the reward system

A main effect of *group* was identified for *FA* (*F*(2,56) = 3.847, *p* = 0.033,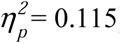). FA significantly decreased linearly from the HC group, to the D- and the D+ group (*t*(59) = −2.264, *p* = 0.027). FA was significantly reduced in the left posterior cingulum subdivision in the D+ group (*t*(59) = 2.673, *p*_*corr*_ = 0.029) and the D-group (*t*(59) = 3.09, *p*_*corr*_ = 0.009) compared to the HC group. Significant *group*tract(FW)* (*F*(8,224) = 2.412, *p* = 0.016, 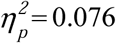) and *group*tract(FW)*hemisphere* (*F*(8,224) = 2.05, *p* = 0.042, 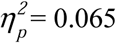) interactions were found, indicating that group differences in FW vary according to tract and hemisphere. FW was significantly increased in the right middle cingulum subdivision in the D-group compared to the HC group (*t*(59) = −3.039, *p*_*corr*_ = 0.011) and in the left MFB in the D+ group compared to the HC group (*t*(59) = −2.594, *p*_*corr*_ = 0.009) (see Figure S4).

### Contribution of global topology and measures in the reward system to depression severity

Global topology measures explained 21.7% 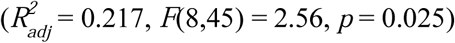 of variance in GDS scores (see Figure 2). *Modularity FW* was a significant independent predictor of GDS scores (*β* = 0.966, *partial r* = 0.269, *p* = 0.05) and *modularity FA* showed a non-significant trend for an independent effect (*β* = −0.866, *partial r* = −0.25, *p* = 0.069).

**Figure 2.**
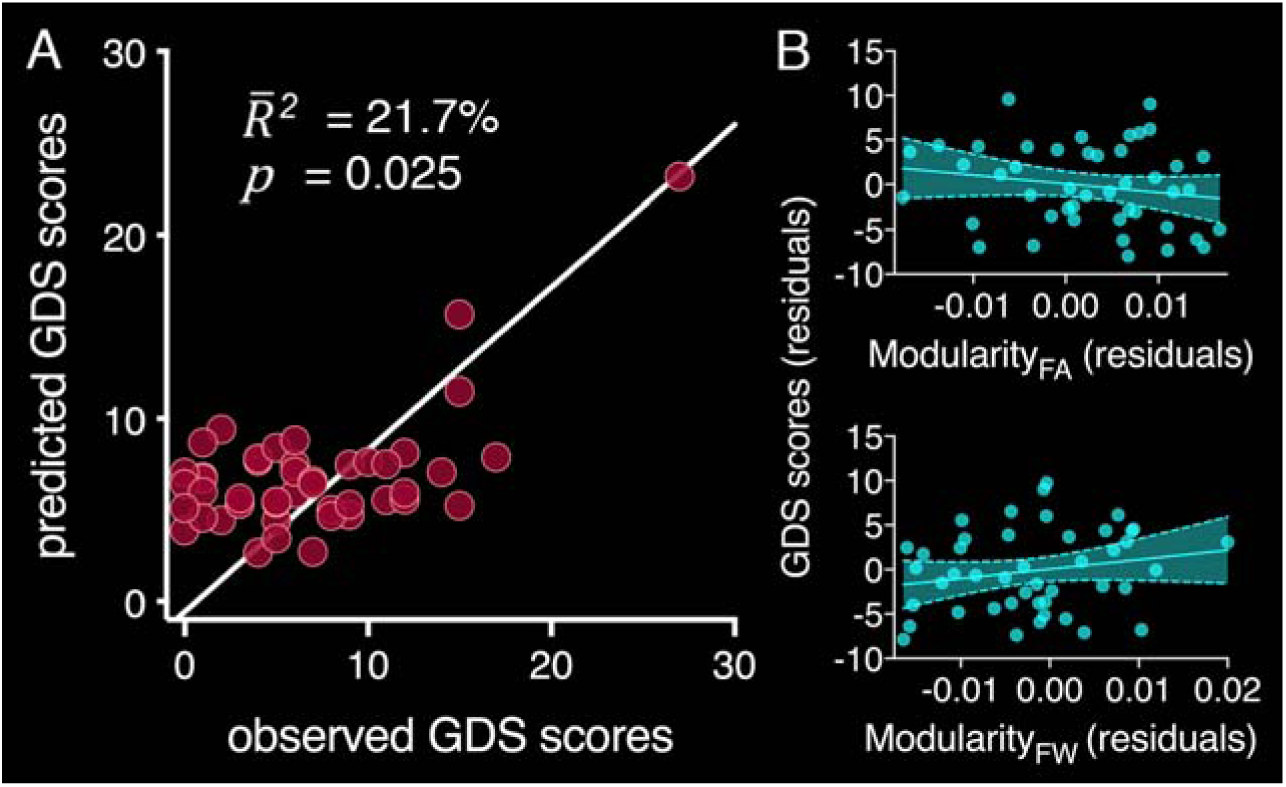
A) Goodness of fit of the regression model with global efficiency (FA/FW), modularity (FA/FW), and centrality coefficient (FA/FW) as predictor variables and GDS scores as outcome variable, including covariates. Observed GDS scores on the x-axis are plotted against predicted GDS scores from the regression model on the y-axis. B) partial correlation plots for the trend-level independent predictor modularity FA (top) and the significant independent predictor modularity FW (bottom).

The final model of a stepwise linear regression analysis with all measures of FA, FW and grey matter volume included 16 variables and explained 76.8% 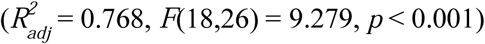 of the variance in GDS scores. FA of the bilateral anterior, middle, posterior and left parahippocampal cingulum subdivisions, as well as FW in the left MFB, right middle and left posterior cingulum subdivisions were significant independent predictors of GDS scores (see Table 2 and Figure 3). Furthermore, the volumes of the left thalamus, bilateral amygdala, right nucleus accumbens and bilateral orbitofrontal cortex were also significant independent predictors of GDS scores.

**Table 2.** Regression analysis - significant independent predictors of GDS scores

|  |  |  |  |  |
| --- | --- | --- | --- | --- |
| predictor variable | standardized $\beta$ | t-statistic | partial r | p-value |
| *fractional anisotropy* |  |  |  |  |
| left anterior cingulum | 0.33 | 2.81 | 0.48 | 0.009 |
| right anterior cingulum | 0.30 | 2.89 | 0.49 | 0.007 |
| left middle cingulum | -0.34 | -2.83 | -0.48 | 0.009 |
| right middle cingulum | -0.27 | -2.35 | -0.41 | 0.026 |
| left posterior cingulum | -0.59 | -5.55 | -0.73 | <0.001 |
| right posterior cingulum | -0.69 | -6.02 | -0.76 | <0.001 |
| left parahippocampal cingulum | -0.45 | -4.02 | -0.61 | <0.001 |
| *free-water* |  |  |  |  |
| right middle cingulum | 0.55 | 4.81 | 0.68 | <0.001 |
| left posterior cingulum | 0.35 | 3.23 | 0.53 | 0.003 |
| left medial forebrain bundle | 0.67 | 6.48 | 0.78 | <0.001 |
| *grey matter volume* |  |  |  |  |
| left thalamus | -0.59 | -5.35 | -0.72 | <0.001 |
| left amygdala | -0.26 | -2.39 | -0.42 | 0.024 |
| right amygdala | 0.28 | 2.59 | 0.45 | 0.02 |
| right nucleus accumbens | 0.36 | 2.86 | 0.48 | 0.008 |
| left orbitofrontal cortex | 0.69 | 3.91 | 0.60 | 0.001 |
| right orbitofrontal cortex | -0.72 | -3.82 | -0.59 | 0.001 |

*Note.* GDS = geriatric depression scale

**Figure 3.**
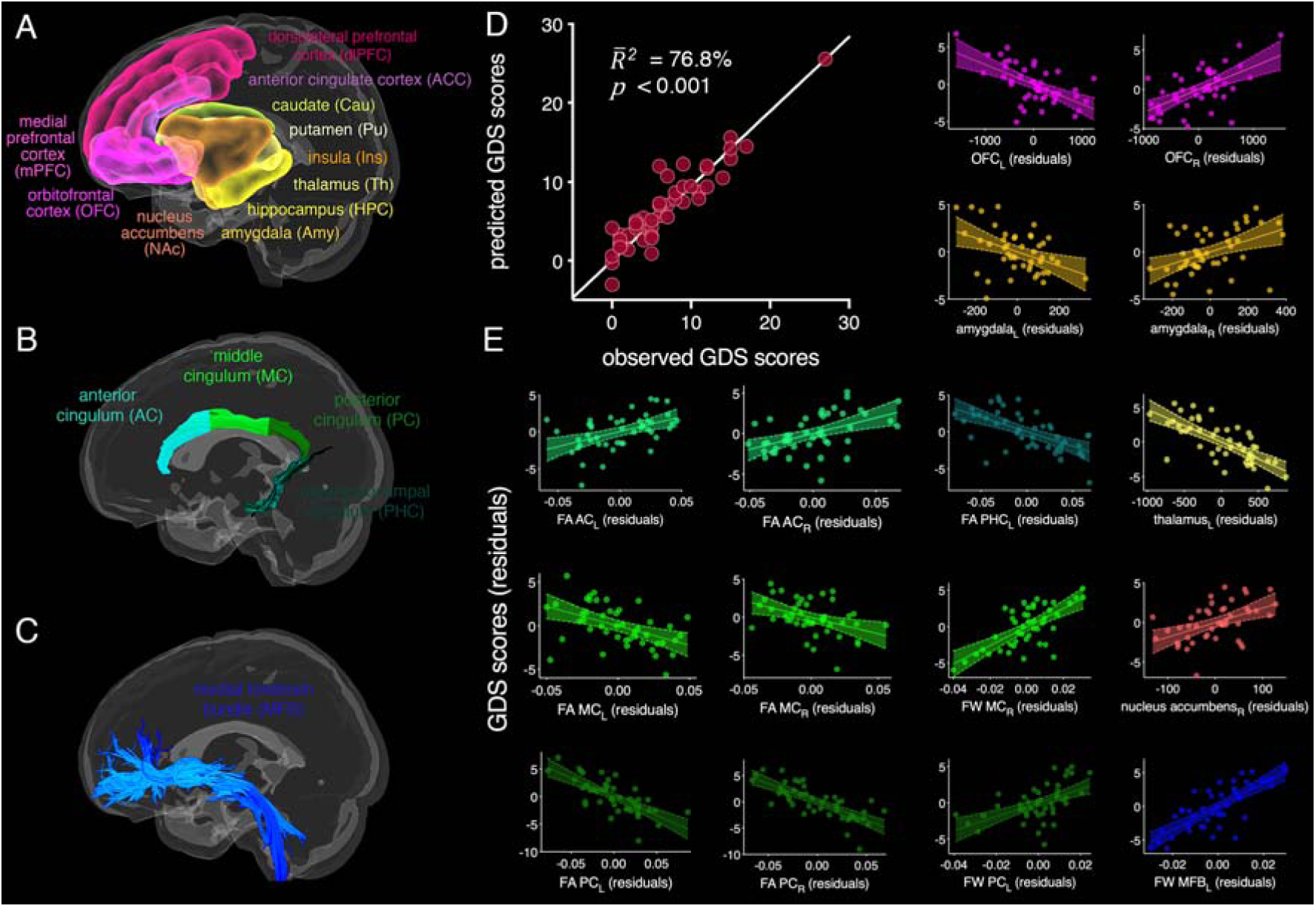
A) Grey matter structures of the reward system B) Cingulum bundle subdivisions from one representative participant C) Medial forebrain bundle (MFB) from one representative participant D) Goodness of fit of the regression model with FA/FW and grey matter volume as predictor variables and GDS scores as outcome variable, including covariates. Observed GDS scores on the x-axis are plotted against predicted GDS scores from the regression model on the y-axis. E) partial correlation plots for the significant independent predictors of GDS scores.

## Discussion

In this study, we found PSD to be associated with global network topology and subnetworks identified from structural connectome analysis that mapped predominantly onto fronto-subcortical regions and connections defined as constituting the reward system. Our focused analysis of grey and white matter correlates within the reward system showed that grey matter volumes of this circuit, together with FA and extracellular FW content of major connection pathways in this system were collectively predictive of PSD severity.

Specifically, the global graph theoretical measure modularity calculated from FW increased with depression severity and subnetworks identified by structural connectome analyses were based on measures of white matter microstructure and FW volume in patients with PSD relative to healthy controls. These predominantly intrinsic frontal and fronto-subcortical subnetworks are typically disrupted in major depressive disorder (MDD). Our findings indicate that the structural basis of PSD, like MDD in the absence of stroke, resides in brain circuits typically involved in motivation, emotions and memory.^16^ In PSD, these alterations may be remote from the infarct itself. Furthermore, to the extent that enhanced FW in the extracellular space is indicative of inflammatory processes, our findings suggest that neuroinflammation may contribute to the development of PSD.

Graph theoretical analyses found that increasing modularity estimated from FW, predicted depression severity. Modularity indicates that nodes are forming communities via dense connections to one another, which have only sparse long-range connections to nodes from other modules. A recent study in individuals with treatment resistant depression found that noninvasive neurostimulation led to a significant reduction of depressive symptoms over time, which was associated with decreased modularity.^30^ The authors concluded that transient changes in modular network configuration may be required to alleviate depressive symptoms. To the extent that reduced FA with a simultaneous increase in extracellular FW volume has previously been interpreted to indicate neuroinflammation,^31^ our finding may suggest that modules of densely packed neuroinflammation across the brain are linked to the development of PSD, possibly in conjunction with compromise of long-distance connections.

On closer examination of structures in the reward system, we observed structural changes in this circuit that account for much of the variability in depression after stroke. The origin of these observed changes, however, is unclear. Several explanations are plausible. (1) It is conceivable that microstructural changes in the reward system represent an underlying liability for depression and that events such as a stroke, lead to the development of depressive symptoms. This is not the same as pre-existing depression. None of the participants reported pre-stroke mood symptoms. (2) The observed changes in the reward system may be secondary to infarction elsewhere in the brain through neuronal degeneration distal to the lesion or neuroinflammatory processes along descending white matter pathways. (3) Infarction and changes in the reward system may share a common causation, such as underlying vascular disease.

The grey matter volume changes and white matter alterations observed in our study closely resemble structural changes reported in MDD. Interestingly, family studies suggest that microstructural changes in the cingulum bundle may represent a biomarker of vulnerability for MDD.^32^ Furthermore, structural and functional abnormalities in the amygdala, nucleus accumbens, thalamus and orbitofrontal cortex (see Table S1) have been reported in MDD and also in healthy individuals with elevated levels of depressive symptoms^33^ and familial risk of depression.^34^ Taken together, these findings may suggest that individuals with altered cingulum bundle microstructure and grey matter changes in the reward system are already at increased risk for MDD and that the event of a stroke increases their chance of developing depressive symptoms.^33^ This might also explain why, similar to previous studies, we were unable to identify strong associations between lesion locations and PSD. Rather than being caused by injury to specific brain regions, it is possible that premorbid microstructural changes in the reward system may render some individuals more susceptible to develop PSD than others in the context of any given lesion.

Our global, whole-brain connectome approach and the localized investigation of the reward system both identified increased FW volume in fronto-subcortical connections in patients with PSD, but not in patients without PSD, relative to healthy controls. Freely diffusing water molecules are characteristic of cerebrospinal fluid and edema, but can also be indicative of more subtle neuroinflammatory processes.^31^ This is because neuroinflammation increases the fractional volume of water molecules diffusing freely in the interstitial extracellular space, where microglia and other immunoreactive cells mediate immune defense.^31^ Neuroinflammation is induced via the release of pro-inflammatory cytokines in response to psychological stress, such as often preceding the onset of MDD, or physiological insult, as caused by stroke. Stroke elicits a cascade of neuroinflammatory events, which have neuroprotective properties and foster neuroplasticity, but can also cause secondary cell death.^35^ Several regions in the reward system have been reported to be specifically vulnerable to neurotoxic effects of pro-inflammatory cytokines,^36^ which, in turn, deplete serotonin and thereby contributes to the development of depressive symptoms.^37^ Given that the patients in this study were scanned approximately 30-95 days post-stroke, it is conceivable that prolonged neuroinflammation in the reward system is a secondary consequence of lesions elsewhere in the brain.

Stroke is strongly associated with vascular risk factors such as diabetes mellitus, smoking, atrial fibrillation, ischemic heart disease, hypertension and cholesterol levels.^38^ It is possible that vascular risk factors are a common causation of both, infarction and microstructural changes in the reward system. However, we did not observe any associations between vascular risk factors and structural measurements in the reward system, indicating that a common causation seems unlikely. While controlling for the effects of covariates in our analyses, only time since stroke was significantly associated with depressive symptoms, whereby depression severity increased with increasing number of days since stroke. This is particularly important as several neural connections regenerate and rearrange in the weeks to months after stroke. Following stroke survivors longitudinally and investigating microstructural changes in the reward system in the acute phase compared to three to six months post-stroke, when depressive symptoms typically peak^2^ would represent an important avenue for future research.

Estimates of white matter microstructure, grey matter volume and extracellular FW in the reward system were strongly associated with depression severity across the entire spectrum of GDS scores, indicating that its sensitivity is not limited to either high or low depression scores. This is particularly important as the characterization of highly sensitive, non-invasive neuroimaging biomarkers to identify individuals at high-risk for PSD is a necessary first step for the implementation of individualized interventions such as low dose antidepressants, anti-inflammatory medication and other prophylactic treatments to prevent the development of PSD. Interestingly, increasing evidence suggests that antidepressant medications possess anti-inflammatory properties,^39^ which may aid the recovery of structural damage restore serotonergic activity.

The present study had several limitations. Our finding of increased FA in the anterior middle cingulum might be biased by crossing projections in this area from the internal capsule. FA has previously been observed to be increased in the internal capsule^40^ of patients with MDD. Future studies, using multi-shell diffusion MRI sequences may wish to investigate the crossing fiber populations in this region. Although the sample size was adequate to assess associations with sensitive and quantitative microstructural measures, it was insufficient for definitive voxel-wise analyses such as VLSM. However, previous VLSM studies^8-10^ and meta-analyses on lesion location^13,14^ have included large numbers of patients and have still been unable to identify brain regions specific to PSD. The purpose of our analysis of lesions was largely to show consistency with these studies and that there was no strong lesion effect arising as an idiosyncrasy of this sample.

In summary, we found global and local structural brain changes to be associated with PSD. Specifically, PSD was strongly associated with white and grey matter measurements in the reward system, similar to those observed in MDD and independent of lesion location.

Evidence of increased extracellular FW in the reward system might indicate a role for neuroinflammation in the development of PSD. Evaluation of structure within this system presents the opportunity to define biomarkers, which could identify individuals at high-risk for developing PSD, who might benefit from early interventions to prevent the development of depressive symptoms.

## Data Availability

This dataset is not publicly available

## Acknowledgements

This study was funded by the Medical Research Council, UK (grant reference MR/K022113/1) and the European Commission Horizon 2020 Health Programme (CoSTREAM, grant agreement 667375). The authors thank the coordinators of the King’s Hyperacute Stroke Research Centre for their help with identifying and recruiting patients, and the manager and staff of the NIHR Wellcome Trust King’s Clinical Research Facility.

